# rRNA analysis based on long-read high-throughput sequencing reveals a more accurate diagnostic for the bacterial infection of ascites

**DOI:** 10.1101/2020.09.14.20194134

**Authors:** Xiaoling Yu, Wenqian Jiang, Xinhui Huang, Jun Lin, Hanhui Ye, Baorong Liu

## Abstract

Traditional pathogenic diagnosis presents defects such as a low positivity rate, inability to identify uncultured microorganisms, and time-consuming nature. Clinical metagenomics next-generation sequencing can be used to detect any pathogen, compensating for the shortcomings of traditional pathogenic diagnosis. We report third-generation long-read sequencing results and second-generation short-read sequencing results for ascitic fluid from a patient with liver ascites and compared the two types of sequencing results with the results of traditional clinical microbial culture. The distribution of pathogenic microbial species revealed by the two types of sequencing results was quite different, and the third-generation sequencing results were consistent with the results of traditional microbial culture, which can effectively guide subsequent treatment. Short reads, the lack of amplification and enrichment to amplify signals from trace pathogens, and host background noise may be the reasons for high error in the second-generation short-read sequencing results. Therefore, we propose that long-read-based rRNA analysis technology is superior to the short-read shotgun-based metagenomics method in the identification of pathogenic bacteria.

## Introduction

Distinguishing and identifying the microorganisms that cause infection is very important for the treatment and survival of patients. The traditional pathogenic diagnosis method generally involves culture, nucleic acid amplification assays and serological analysis (1) and presents shortcomings such as a low positivity rate, inability to identify uncultured microorganisms, and time-consuming nature. Moreover, the existing molecular tests cannot detect the characteristics of newly evolved genes in pathogens that spread among humans, animals, and the environment(2).

Metagenomics is the application of modern genomics technology to directly study microbial communities in the natural environment without the need for the isolation and laboratory cultivation of individual species (3). Next-generation sequencing (NGS), also known as high-throughput sequencing, allows the simultaneous and independent sequencing of thousands to billions of DNA fragments (4). Clinical metagenomics next-generation sequencing (mNGS) is a method for the comprehensive analysis of microorganisms and host genetic material (DNA and RNA) in specimens from patients (5).

mNGS can be used to detect any pathogen and provides auxiliary genomic information required for evolutionary tracking, strain identification, and drug resistance prediction (4). mNGS has been applied in clinical and public health fields. For example, NGS was used to clinically diagnose the pathogen responsible for meningoencephalitis in a 14-year-old boy with severe combined immunodeficiency (SCID), and a *Leptospira* strain that may cause encephalitis was found (6). Additionally, in the field of public health, researchers used whole-genome sequencing (WGS) to conduct the epidemiological analysis of an *Acinetobacter baumannii* infection that broke out in a British hospital in 2010 (7).

At present, mNGS can be performed with either second-generation sequencing technology or third-generation sequencing technology. The available second-generation short-read sequencing platforms mainly include the Illumina, SOLiD, Ion Torrent and BGISEQ systems, while the third-generation long-read sequencing platforms mainly include the Nanopore and PacBio systems (8). PacBio sequencing is a long-read sequencing platform that uses a single-molecule real-time (SMRT) chip as a synthetic sequencing vector (9). The sequence data generated by PacBio sequencing are helpful for determining the species and genus of pathogenic microorganisms involved in infections and do not require a traditional pathogenic microorganism culture step. Compared with the traditional detection technology for pathogenic microorganisms, this technology presents great advantages in the detection of microorganisms that are difficult to isolate and culture in clinical settings and some uncultured microorganisms. PacBio sequencing has the potential to identify pathogenic microorganisms in the clinical environment, which is helpful for making clinical treatment decisions. Here, we describe the PacBio sequencing results and second-generation Illumina sequencing results for an ascitic specimen from a patient with liver ascites, and we compare the two types of sequencing results with the results of traditional clinical microbial culture to assess the advantages and disadvantages of different sequencing strategies for specific specimen types.

## Methods

### Sample collection and DNA extraction

According to clinical standard operating procedures, the medical staff performed abdominal puncture to collect 3-5 ml of ascites and injected the fluid into a culture bottle (Zhengzhou Antu Biological Co., Ltd., China) for enrichment culture. After the culture was shown to be positive, it was transferred to Columbia blood agar plates (Zhengzhou Antu Biological Co., Ltd., China) for anaerobic culture at 35°C for 48 hours. Subsequently, MALDI-TOF-MS (VITEK MS, BioMerieux SA, BioMerieux Inc., France) (10) was used to identify the cultivated pathogenic microorganisms.

The sodium chloride, Tris-HCl, EDTA (STE) DNA extraction method (11) was used to extract the total DNA of the microorganisms in the ascitic samples. First, 1 mL of ascites was centrifuged at 10,000×g for 1 min, after which the supernatant was discarded, and 1× STE buffer was added to resuspend the pellet. Then, an MP Biomedicals sample preparation instrument was used to homogenize the ascitic specimen with beads, and the DNA of the ascitic specimen was extracted according to STE method. Name the sample as M173251, which was used for subsequent second-and third-generation sequencing.

### Next-generation sequencing and data analysis

We chose the Illumina platform to carry out short-read NGS. A library was constructed using the Hieff NGS Fast Tagment DNA Library Prep Kit for Illumina (YEASEN, China), and the library was sequenced in an Illumina NovaSeq sequencer.

There is an existing mature commercial analysis process for Illumina sequencing (12). High-quality sequencing data were generated by removing low-quality reads, adapter contamination, and duplicated reads using Trim Galore (13). Host contamination present in the sample required the comparison with the host sequence to filter out possible sequences from the host (14–16). Human sequence data were excluded and mapped to a human reference genome (hg19) using Bowtie2 software (parameter settings: --end-to-end, --sensitive, -I 200, -X 400). After removing human sequences, Kraken2 software (v2.0.7-beta, https://www.ccb.jhu.edu/software/kraken2/) was used to compare and annotate the remaining sequences with the MiniKraken2_v2 database, to integrate the comparison results for the sample, and to obtain species abundance statistics. Starting from the relative abundance tables for different classification levels, the top 9 species with the relative abundance in the sample (group) were selected, the remaining species were set as others, and the corresponding species annotation results for the sample were plotted in relative abundance histograms at different classification levels.

### Third-generation sequencing and data analysis

We chose the PacBio platform to carry out the third-generation high-throughput sequencing analysis of full-length rRNA sequences based on long reads. According to the full-length 27F and 1492R primers for 16S rRNA, the specific primers 27F_0074 (5’ <u>*TGACAGTATCACAGTG*</u>AGRGTTTGATYNTGGCTCAG 3’) and 1492R_0074 (5’ <u>*CACTGTGATACTGTCA*</u>TASGGHTACCTTGTTASGACTT 3’) with barcodes were synthesized (where the italicized underlined sequence is the barcode sequence). PCR amplification was carried out, and the products were purified, quantified and homogenized to generate a sequencing library (SMRT Bell). The constructed library was inspected first, and the qualified library was sequenced with PacBio Sequel.

Data preprocessing: After exporting the PacBio offline data to circular consensus sequencing (CCS) files (the CCS sequence was obtained using the Smrtlink tool provided by PacBio), the following three main steps were performed: 1) CCS and to obtain barcode CCS sequence data; 2) CCS length filtering: the barcode CCS was filtered to obtain the effective sequence; and 3) chimera removal: Optimization-CCS sequence was obtained by using UCHIME v4.2 software to identify and remove the chimera sequence. identification: Lima v1.7.0 software was used to identify CCS through barcodes and to obtain barcode CCS sequence data;2)CCS length filtering: the barcode CCS was filtered to obtain the effective sequence; and 3) chimera removal:Optimization-CCS sequence was obtained by using UCHIME v4.2 software to identify and remove the chimera sequence. Information analysis content: divide OTU, diversity and difference analysis. The PacBio sequencing results were clustered with Usearch software (17) at a 97% similarity level to obtain OTUs, and the OTUs were annotated using ribosomal database project (RDP) classifier based on the Silva (bacteria) and UNITE (fungi) taxonomic databases. The comparison of representative OTU sequences with a microbial reference database by RDP classifier provides species classification information corresponding to each OTU, after which the sample community composition can be quantified at each level (phylum, class, order, family, genus, species). QIIME software was used to generate species abundance tables at different taxonomic levels, and R language tools were then used to draw community structure diagrams at each taxonomic level for the sample.

According to the existing taxonomic database of microbial species provided by the National Center for Biotechnology Information (NCBI), the species abundance information obtained by sequencing was regressed to the phylogenetic tree of the database by using MEGAN (18) software. The MEGAN taxonomic tree diagram was then drawn to comprehensively elucidate the evolutionary relationships and differences in abundance among all microorganisms in the samples according to the entire classification system.

## Results

Hundreds of single colonies were obtained through clinical bacterial culture, and the morphology of the colonies was similar. A single colony was selected to identify the cultivated pathogenic microorganism via MALDI-TOF-MS, resulting in its identification as *Clostridium difficile (C. difficile)*.

Illumina sequencing generated 15,636,287 reads, each of which was 150 bp in length, and 637,196 reads remained after filtering out the host sequences. We selected the top nine species in terms of abundance after removing the host sequences, merged the remaining species into a group of others, and drew the species distribution map for the sample at the species level (Figure 1A). With the exception of the others group (40%), *Staphylococcus aureus (S. aureus)* showed the highest proportion at close to 40%, followed by *Klebsiella pneumoniae* at 22%, *Burkholderia pseudomallei* at approximately 14%, and *Aeromonas hydrophila* at approximately 7%. Small contributions of *Pasteurella multocida, Campylobacter jejuni, Mycoplasma mycoides, Polynucleobacter necessarius* and *Whitefly endosymbionts* were also found, whereas *C. difficile* was not.

**Figure 1.**
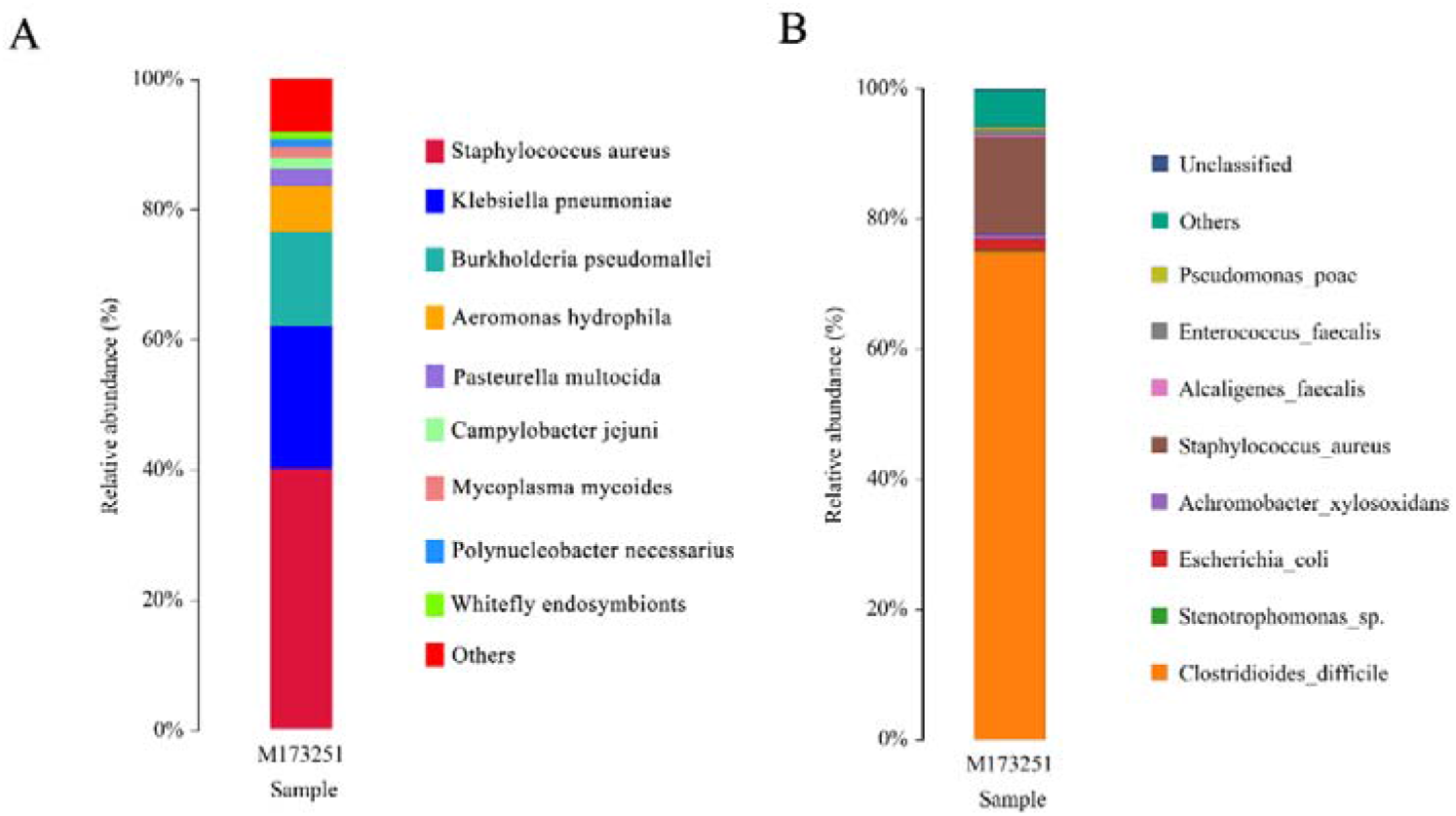
Statistical charts of the species annotation results of next-generation sequencing and PacBio sequencing at the species level. Each color represents a species, and the length of the color block represents the relative abundance ratio of the species; unclassified represents species that have not been taxonomically annotated.

Figure 1A. Statistical chart of species annotation results of next-generation sequencing (Illumina) at the species level.

Figure 1B. Statistical chart of species annotation results of PacBio sequencing at the species level.

A total of 9385 CCS sequences were obtained from PacBio sequencing after identification based on barcodes. The number of sample sequences at each stage was processed according to the statistical data to evaluate data quality. The evaluation results for the sequencing data of the sample are shown in Table 1. We filtered out sequences other than those of 1 kb-1.8 kb in length and obtained 8702 valid CCS sequences, among which 8562 CCS sequences were used for subsequent analysis, which accounted for 91.23% of the CCS reads obtained by sequencing. After statistical quality control and filtering, the number of sequences of reads in the corresponding length range in the sample was plotted in an effective tag length distribution map (Figure 2), and most of the sequences were distributed between lengths of 1400 bp and 1500 bp.

**Table 1.**
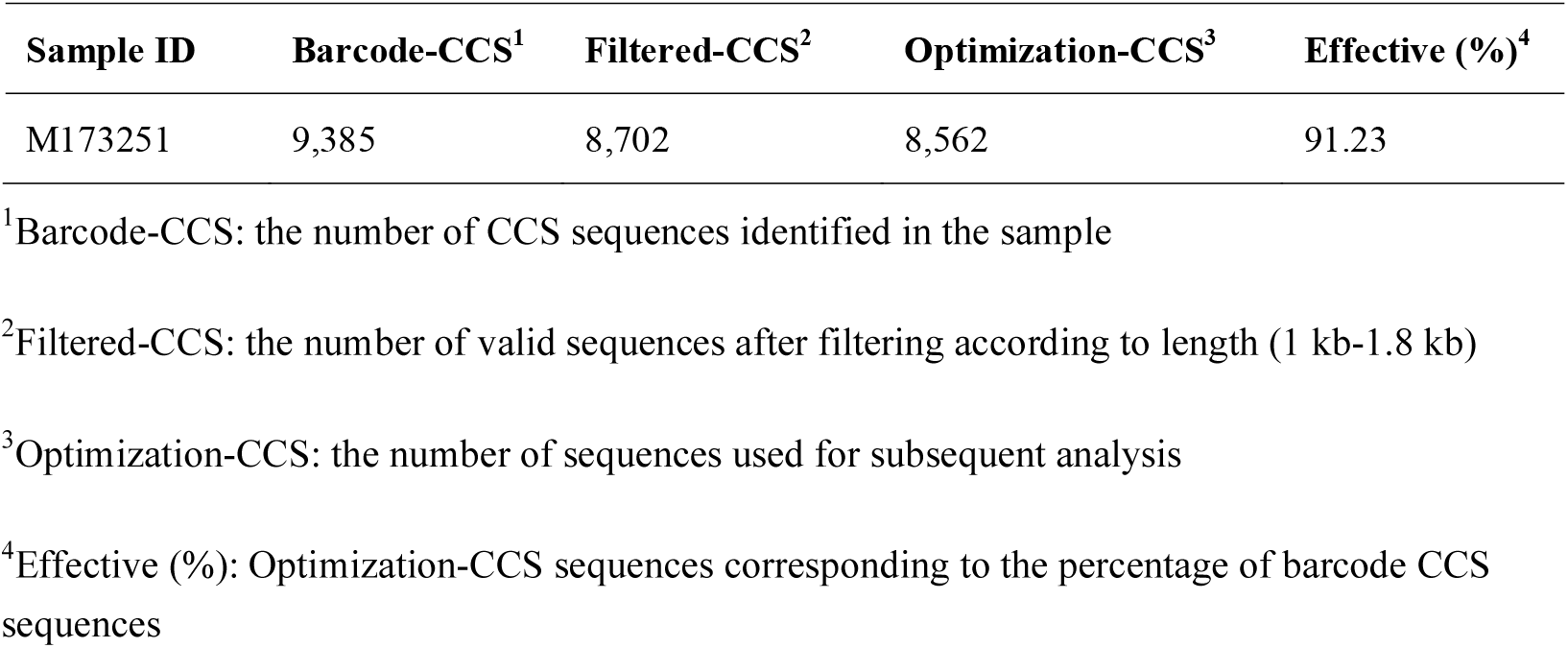
Statistics of sample sequencing data processing results

**Figure 2.**
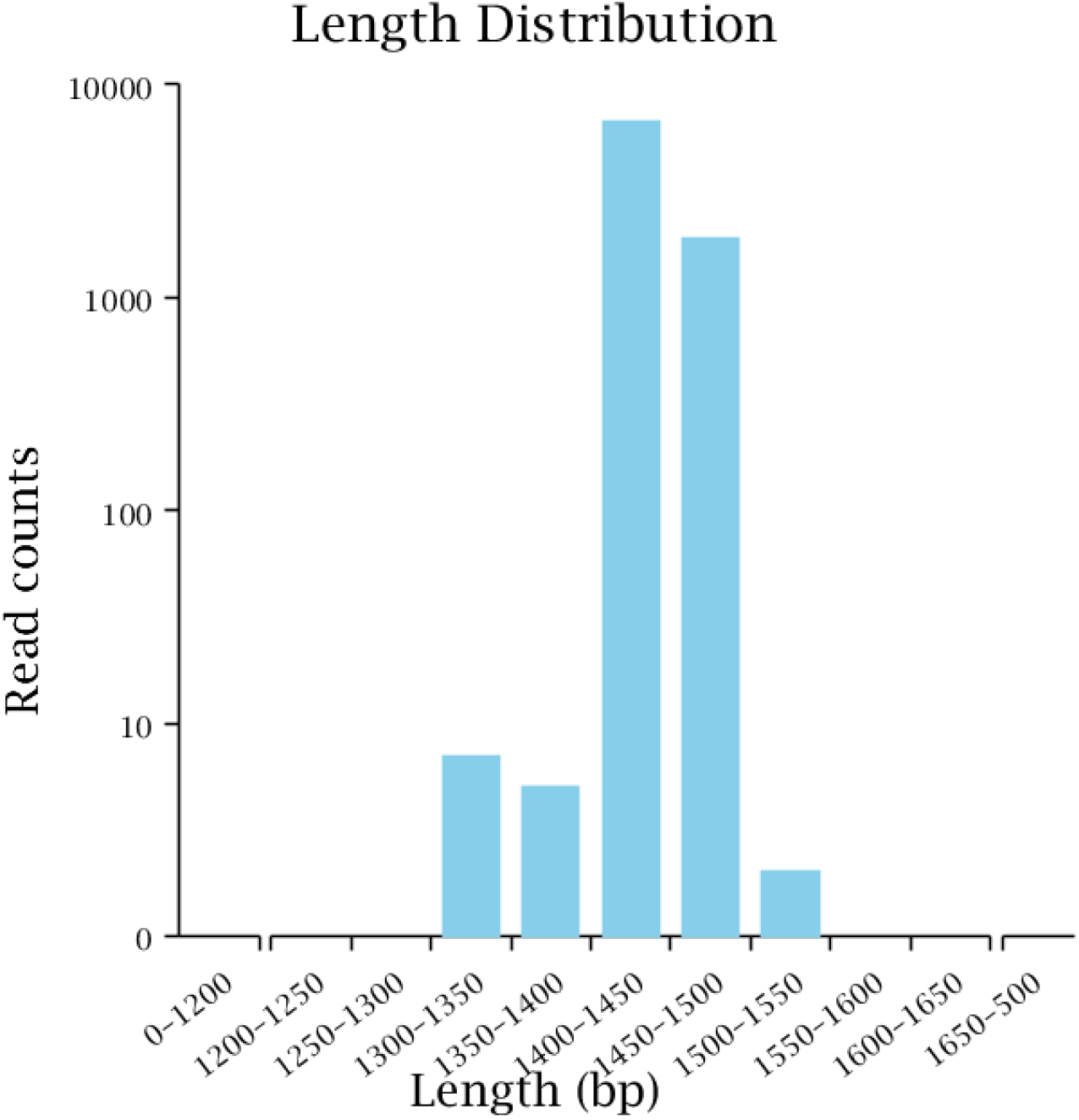
Effective tag length distribution diagram. The abscissa represents the length range, and the ordinate represents the number of reads.

The sequencing results identified 31 OTUs through clustering. We selected the top nine species according to abundance and merged the remaining species into a group of others, after which the species distribution map of the sample was drawn at the species level (Figure 1B). *C. difficile* showed the highest proportion, of approximately 73%, followed by *S. aureus*, at approximately 14%, *Escherichia coli (E. coli)*, at approximately 2%, *Achromobacter xylosoxidans (A. xylosoxidans)*, at approximately 1%, and the others group, accounting for approximately 6%; small contributions of *Stenotrophomonas sp., Pseudomonas poae, Alcaligenes faecalis* and unclassified microbes were also found.

A MEGAN taxonomy dendrogram was drawn on the basis of the PacBio sequencing results. The MEGAN taxonomy dendrogram (Figure 3) shows the predominance of the *Peptostreptococcaceae* (to which *C. difficile* belongs) and *Staphylococcus* groups, which was consistent with the results of cluster analysis.

**Figure 3.**
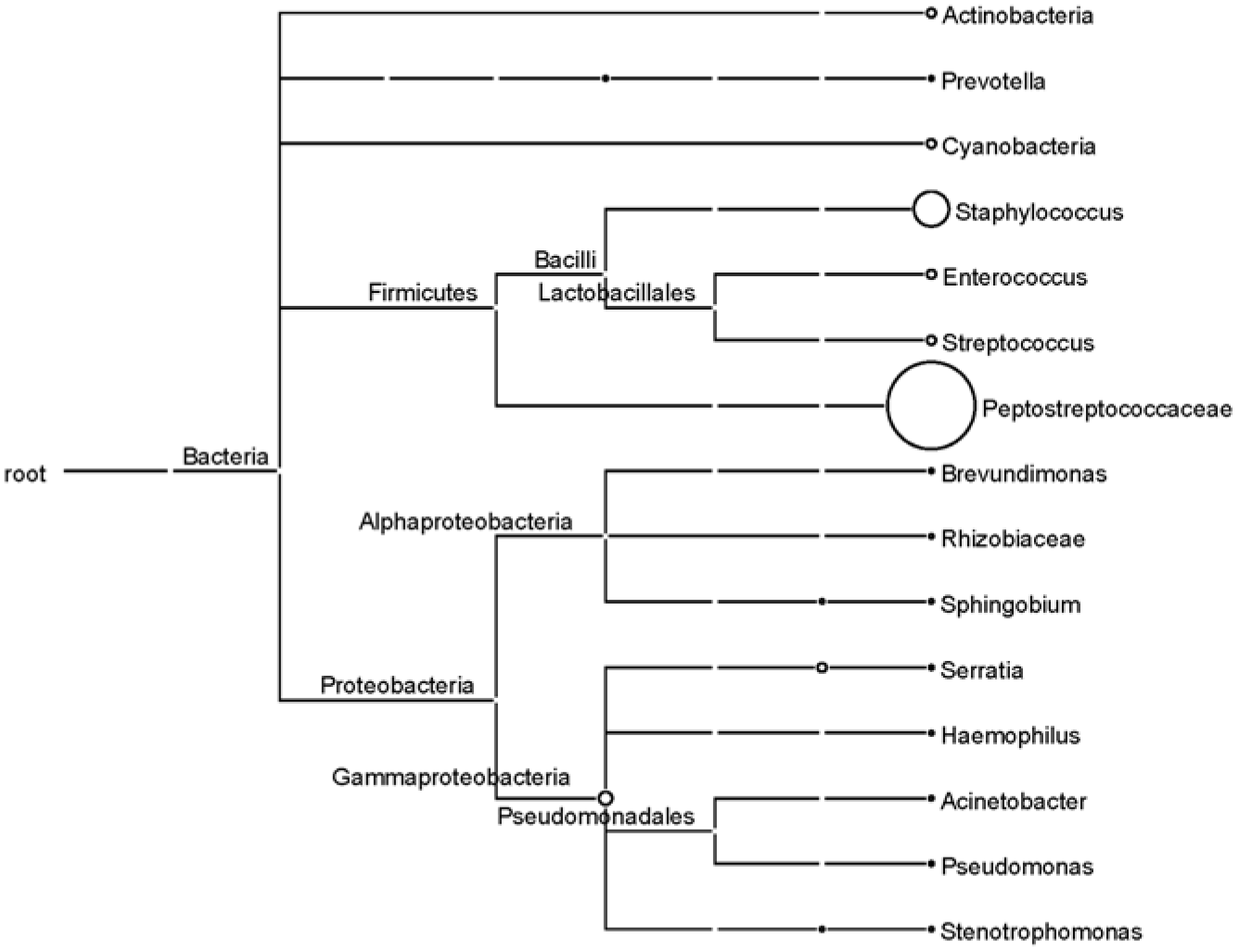
Phylogenetic diversity of the ascitic sample sequences calculated by MEGAN on the basis of PacBio sequencing. Each circle represents a taxon in the NCBI taxonomy and is labeled with its name and the number of reads that were either directly assigned to the taxon or indirectly assigned via one of its subtaxa. The size of the circles is logarithmically scaled to represent the number of reads assigned directly to the taxon (18).

## Discussion

*C. difficile* is an obligate anaerobic gram-positive bacillus that is ubiquitous in nature and is transmitted orally through feces (19). *C. difficile* produces metabolically inactive endospores, making it is resistant to gastric acid and most kinds of antibiotics (19). *C. difficile* can produce two toxins: enterotoxin A and cytotoxin B (20). The pathogenicity of *C. difficile* is based on the fact that at least one of the two toxins acts as a glycosyltransferase to modify guanosine triphosphatases in intestinal epithelial cells and cause the disruption of the actin cytoskeleton (21). *Clostridioides difficile* infection (CDI) is a symptomatic infection caused by the spore-forming bacterium *C. difficile*, and it is one of the common infections found in hospitals (22). The symptoms of CDI include watery diarrhea, fever, loss of appetite, nausea, and abdominal pain, which may lead to colitis in severe cases (23). The complications of CDI include dehydration, low blood pressure, electrolyte imbalances, and more serious problems such as bowel perforation, kidney failure, or even death (23). CDI is a common cause of diarrhea. Among all cases of antibiotic-related diarrhea, a total of 15%-25% are caused by CDI (24). Because *C. difficile* is an obligate anaerobe, it is difficult to isolate and cultivate in a clinical setting. The current methods for the clinical diagnosis of CDI include nuclear acid amplification tests (NAAT), toxigenic *C. difficile* culture, enzyme immunoassays (EIA) for the A1B toxin and glutamate dehydrogenase (GDH), and the *C. difficile* cytotoxin neutralization assay (19).

The medical history of the patient is complicated. He suffered from primary liver cancer, hepatitis B, and cirrhosis. He was hospitalized again for hepatic ascites and abdominal cavity infection. *C. difficile* was cultured from his ascitic fluid, and the third-generation sequencing results also indicated the occurrence of CDI. Mild to moderate CDIs are treated with metronidazole, and for severe infections, vancomycin is administered orally (25). Although only *C. difficile* was cultured from the ascites of this patient, the abdominal infection was indicated to be a compound infection. The PacBio sequencing results also showed that in addition to *C. difficile*, the ascites contained *S. aureus, E. coli, A. xylosoxidans* and other microorganisms. Infections caused by methicillin-susceptible *S. aureus* (MSSA) are usually treated with β-lactam antibiotics, such as cefazolin; however, methicillin-resistant *S. aureus* (MRSA) infections show resistance to penicillin, oxacillin and other β-lactam antibotics, so it is necessary to use more effective drugs such as linezolid against gram-positive bacteria (26). *A. xylosoxidans* is an aerobic gram-negative bacillus that is widely distributed in the environment and mainly causes health care-associated infections (27). Most of the isolates of this species show in vitro sensitivity to carbapenems and piperacillin/tazobactam (28). After the patient was administered metronidazole, cefoperazone, sulbactam, moxifloxacin, linezolid, and biapenem for anti-infection treatment, his symptoms improved significantly. Metronidazole is used to treat *C. difficile*, cefoperazone, sulbactam, biapenem, and moxifloxacin and exerts obvious antibacterial activity against *E. coli* and *A. xylosoxidans*, while linezolid is used to treat infections caused by MRSA.

Clinical bacterial culture resulted in the cultivation of only *C. difficile*. Both the Illumina and PacBio sequencing results showed that the patient’s ascites contained more than one pathogenic microorganism, but the species distribution indicated by the second- and third-generation sequencing results was significantly different. The second-generation sequencing results showed that *S. aureus* accounted for the highest proportion (approximately 40%) of the sample in the species distribution map at the species level (Figure 1A), while *C. difficile* was not detected. In contrast,the third-generation sequencing results showed that *C. difficile* accounted for approximately 73% of the species distribution map at the species level, followed by *S. aureus*, accounting for approximately 14% (Figure 1B). The microbial nucleic acids of the sample are dominated by background human host sequences in Illumina sequencing, where the vast majority of reads (usually >99%) come from human hosts (4), while relatively few microbial reads are obtained. The rRNA long-read sequencing method used in this study targets and amplifies 16S rRNA gene sequences in the sample before sequencing, and there is no background noise from the human host. First, the 16S rRNA gene sequence in the sample is amplified to achieve signal amplification, which is more conducive to the detection of some less-abundant microorganisms. In contrast, library construction is performed directly without amplification in Illumina sequencing, which may mean that some low-abundance microorganisms are not detected. More importantly, sequences from different microbial species are preferentially targeted when the second-generation shotgun macrogene library is constructed. For example, the secondary structure of some microorganisms is complex, and the GC content may be high. PCR amplification is also required for the construction of the library, and the same amplification efficiency is not achieved for these sequences as for other microorganism sequences during library construction. Even under PCR-free second-generation sequencing library construction, due to the existence of special structures, preferences will arise in steps such as the addition of adapters. This is not the case for full-length rRNA third-generation sequencing because rRNA is quite conserved. When building a library for third-generation sequencing, the universal rRNA sequencing primers that are used are also fixed and conserved. Most of the rRNA sequences of different bacteria are the same, and only the sequences of variable regions are different. Therefore, compared with second-generation sequencing, it is easier to obtain consistent amplification efficiency under this method. The result of PacBio sequencing is a full-length sequence of the 16S rRNA gene, while the reads of Illumina sequencing are only 150 bp in length, meaning that long DNA strands are interrupted. After sequencing is complete, splicing and assembly are performed, which may also cause errors. It is worth mentioning that to obtain results in the shortest amount of time, the reads used in the current commercial and clinical trials of mNGS for clinical microbiological detection are generally only 50-75 bp in length, which is much shorter than the 150 bp reads used in this study. All of these factors may limit the sensitivity of Illumina sequencing for pathogen detection, resulting in differences in the microbial species and contents identified between Illumina and PacBio sequencing results.

PacBio sequencing does not require the interruption of the genome and can produce longer reads (29), while second-generation sequencing interrupts long DNA strands during library construction, and after sequencing, the reads need to be spliced and assembled. At present, the reads generated by mainstream second-generation sequencers are generally 50-150 bp in length (8). Nanopore sequencers are another third-generation long-read technology that can identify DNA or RNA sequences by detecting the characteristics of current signals caused by charged biomolecules (such as DNA or RNA) through nanopores (30), among which the MinION platform is most commonly used. Compared with PacBio and Illumina sequencing, the accuracy of Nanopore sequencing is much lower, with an error rate of 15% to 40%, and sequencing errors are mainly caused by the insertion and deletion of bases (31). When there are fewer consecutive identical bases in a DNA sequence, the number of bases recognized by nanopore sequencing may produce errors, which is one of the reasons for the low accuracy of this approach.

We compared Illumina NovaSeq sequencing, PacBio sequencing and Nanopore sequencing (Table 2). PacBio is superior to NovaSeq and Nanopore in terms of sequencing time, reading length, accuracy and cost.

**Table 2.** Comparison of Illumina NovaSeq, PacBio and Nanopore sequencing

| Sequencing platform | Time of library preparation | Time of sequencing | Costs/GB (local price) | Data/per person (GB) | Library construction cost | Accuracy |
| --- | --- | --- | --- | --- | --- | --- |
| Illumina NovaSeq (32) | 3-5 hours | Up to 44 hours | \$ 7.14 | 5-8 G | \$ 28.53 | 99.9% |
| PacBio (33) | <3 hours | 0.5 to 30 hours | \$ 9.98 | 29 M | \$ 35.66 | > 99.999% (CCS) |
| Nanopore MinION (31) | 15 min + | Up to 48 hours | \$ 42.43 | No relevant information was found | \$ 141.37 | 60% - 85% |

The research results obtained via the 16S rRNA PCR approach and NGS (Illumina HiSeq) for the ascitic microbiota have been compared, and it was found that whole-genome shotgun-based NGS on the Illumina platform is more suitable for describing the microbiome of ascitic fluid or other low-abundance bacterial DNA samples than the 16S rRNA PCR approach based on the Illumina platform (34). To our knowledge, this is the first study to apply PacBio sequencing to explore the ascitic microbiome. Our experimental results showed that 16S rRNA sequencing based on the PacBio platform was more accurate than mNGS based on the Illumina platform; thus, the accuracy of the sequencing results was ranked as follows from highest to lowest: 16S rRNA sequencing (PacBio), mNGS (Illumina), and 16S rRNA sequencing (Illumina).

## Conclusion

Compared with third-generation long-read sequencing technology, second-generation metagenomic DNA sequencing technology produces shorter reads, and it is difficult to obtain information such as the full-length sequences of the drug resistance genes of pathogenic microorganisms via this approach; furthermore, it may introduce bias and shows low detection rates of some low-abundance intracellular bacteria and fungi with thick cell walls, long sequencing times and other defects [9]. Long-read PacBio sequencing exhibits a higher accuracy rate than second-generation sequencing. Although the sequencing cost per GB of data is higher than that of Illumina sequencing, the amount of data required for pathogenic microorganism sequencing is much lower, and the overall cost is also lower than that associated with Illumina sequencing. In addition, the amount of third-generation full-length rRNA data obtained via the method used in this research is small. Hence, compared with second-generation short reads, the dependence of the subsequent analysis on high-performance computers is low; few calculations are required; and the analysis results can be obtained faster. In contrast, traditional bacterial culture takes a long time, and many pathogenic microorganisms are not easy to cultivate. For example, only *C. difficile* was cultivated from the compound infection of the patient examined in the present study. As PacBio sequencing technology continues to mature, the cost of sequencing will decrease, and the accuracy will be further improved. PacBio sequencing has the potential to become another diagnostic method for pathogenic microorganism detection in the clinical environment.

## Data Availability

All raw data was uploaded to the NCBI-SRA database under the accession number of PRJNA661722.

https://www.ncbi.nlm.nih.gov/sra/PRJNA661722

## Author Approvals

All authors have seen and approved the manuscript, and that it hasn’t been accepted or published elsewhere.

## Ethics approval

All authors ensure that the work described has been carried out in accordance with The Code of Ethics of the World Medical Association (Declaration of Helsinki) for experiments involving humans. The ethical approval of this study was provided by the Institutional Ethical Committee of the Faculty of Medicine, Mengchao Hepatobiliary Hospital of Fujian Medical University, Fujian, China (No. 2019_057_01). Informed consent was obtained for experimentation with human subjects.

## Data Accessibility

All raw data was uploaded to the NCBI-SRA database under the accession number of PRJNA661722 (https://www.ncbi.nlm.nih.gov/sra/PRJNA661722).

